# Molecular diagnosis of Cytomegalovirus infection: clinical performance of Transcription Mediated Amplification (TMA) towards conventional qPCR chemistry on whole blood samples

**DOI:** 10.1101/2023.07.19.23292864

**Authors:** Paolo Bottino, Lisa Pastrone, Elisa Zanotto, Francesca Sidoti, Cristina Costa, Rossana Cavallo

## Abstract

Human Cytomegalovirus (HCMV) infection represents a life-threating pathogen for immunocompromised patients. Molecular quantitative testing on whole blood or plasma represents the gold standard for diagnosis of invasive HCMV infection and for monitoring antiviral treatment in individuals at risk of CMV disease. For these reasons, accurate standardization towards the 1st WHO International standard between different centres and diagnostic kits represent an effort for a better clinical management of CMV-positive patients. Herein, we evaluate for the first time the performance of a new TMA (Transcription Mediated Amplification) kit towards qPCR chemistry, used as routine method, on whole blood samples. 755 clinical whole blood specimens were collected and simultaneously tested with TMA and qPCR assays. Data showed 99.27% agreement for positive quantified samples and 89.39% agreement for those not detected between two tested methods. Evaluation of viraemia in positive samples highlighted a good correlation for TMA and qPCR chemistries in terms of International Units (ΔLog_10_ IU/ml: − 0.29 ± 0.40). TMA assay showed a significant correlation with qPCR also in monitored patients until three months, thus allowing accurate evaluation of viraemia in transplanted patients. Moreover, preliminary data about analytical sensitivity of TMA chemistry onto DBS samples showed 86.54% correlation with whole blood specimens. Thus, TMA chemistry showed a good agreement with qPCR assay, used as current diagnostic routine, and offers important advantages: FDA and IVD approval on plasma and whole blood, automated workflow with minimal hands-on time, random access loading, thus enabling a rapid and reliable diagnostics in HCMV-infected patients.

## Introduction

Human Cytomegalovirus (HCMV), also known as human herpesvirus 5 (HHV-5), is the prototype member of the *Betaherpesvirinae* and the largest member of the virus family *Herpesviridae* (1). It is a ubiquitous virus that infects almost all humans at some time in their lives. The virus was first isolated by three different groups of researchers: Rowe and colleagues, Weller and colleagues, and Smith simultaneously in 1956 (2). The genome is a linear, double-stranded DNA molecule with a 236 kbp size and consisting of two unique regions, each flanked by inverted repeats (3). Despite enclosing a much larger genome, the size of the HCMV capsid is similar to that of other herpesviruses, structured as an icosahedrally ordered nucleocapsid with triangulation number (T) = 16, composed of 12 pentons, 150 hexons, and 320 triplexes (4, 5). Externally to the capsid, it is located the tegument, generally thought to be unstructured and amorphous in nature. However, some structuring is seen with the binding of tegument proteins to the protein capsid. The tegument proteins comprise more than half of the total proteins found within infectious virions (6, 7). Finally, a lipidic bilayer envelope membrane, containing several virus-specific glycoproteins, in turn covers tegument and HCMV nucleocapsid. This envelope is similar in structure and composition to host cell membranes (8, 9). HCMV is a global herpesvirus highly prevalent worldwide with a prevalence of about 100% in both Africa and Asia, and 45.6-95.7% in Europe and North America (10). The heterogeneous CMV seroprevalence appear to be related with race, ethnicity, socioeconomic status and education level (11). Focusing on Italy, the seroprevalence of HCMV appear to be as high as 70–80% (12, 13).

Cytomegalovirus infection can occur during pregnancy and alongside the entire lifetime after delivery, by several transmission routes. As congenital HCMV infection (cHCMV), maternal primary and non-primary infection (exogenous reinfection with a different strain or endogenous viral reactivation) of virus during pregnancy can result in in utero transmission to the fetus (vertical route). Approximately 11% of live-born infants with cHCMV show abnormal clinical signs at birth: hematological disorders, cerebral malformation, chorioretinitis and sensorineural hearing loss (SNHL), the most common sequela (14, 15). Infants without symptoms at birth are also reported to be at risk of developing long-term hearing loss (16).

In the postnatal period, primary HCMV infections are acquired via several ways through contact with infected fluids (e.g., saliva, breast milk, blood products), at home and in nursery schools as community exposure (17). Breastfeeding is known to be the first close contact with the greatest impact, probably due to viral reactivation in the mammary glands and subsequent excretion of Cytomegalovirus into the milk without clinical or laboratory signs of systemic infection (negative serum IgM, negative viraemia) (18). During childhood and early adulthood, HCMV is transmitted through exposure to saliva, stool and urine (10).

Among adults genital secretions are a common fluid for CMV shedding, consistent with several studies that identified sexual risk factors for CMV seropositivity or seroconversion (10, 13, 19). Another important transmission route for primary infection consists in solid organ transplantation (SOT), especially in cases where there is a serological mismatch between donor and recipient (the recipient is CMV seronegative and the donor is seropositive) (20). Otherwise, infection can occur as reactivation in those patients with risk factors such as intense immunosuppression, use of lymphocyte-depleting antibodies or prednisolone, acute rejection, older age in the donor and/or recipient, concomitant viral infections or genetic polymorphisms (21, 22).

HCMV infection is usually asymptomatic in immunocompetent people, although clinical symptoms of primary infection may include a nonspecific glandular fever (mononucleosis) syndrome characterized by flu-like symptoms (23). Instead, in immunocompromised or transplanted patients, HCMV primary infection or reactivation represent a fearsome complication resulting in a viral syndrome, characterized by fever and malaise as well as leukopenia, thrombocytopenia and elevated liver enzymes. Pneumonia, hepatitis, meningoencephalitis, pancreatitis or myocarditis may rarely be present, requiring admission to an intensive care unit (20, 24). HCMV infection may also have indirect effects on graft dysfunction, acceleration of coronary artery atherosclerosis, renal artery stenosis and the emergence of other opportunistic infections (25, 26).

Alongside serological tests and pp65 antigenemia, direct detection of HCMV DNA in clinical specimens is currently the standard method for the diagnosis of HCMV infection (2, 27). Particularly, Quantitative Polymerase chain reaction (qPCR) represents the gold standard and several IVD kits based on HCMV conserved regions were developed in order to detect and quantify HCMV DNA (28–32). Whole blood and plasma are the most common specimens for HCMV qPCR (33–35), although cerebrospinal fluid (CSF) and bronchoalveolar lavage fluid (BAL) are sometimes used (36). Scheduled monitoring of HCMV viremia in immunocompromised individuals is critical for identifying patients at risk for CMV disease, evaluating preemptive therapy and determining treatment response (2).

In addition to qPCR, other molecular techniques such as TMA (Transcription Mediated Amplification) have recently been developed and approved as commercial IVD kits. However, their clinical performance, especially on whole blood, is still under study.

Starting from this perspective, the main aim of our work was to evaluate the clinical performances of TMA chemistry on whole blood samples in a cohort of adult and pediatric individuals. Furthermore, focusing on transplanted patients, HCMV viremia was monitored over time with both chemistries. Finally, for an in-depth evaluation of whole blood matrix, a preliminary analysis was also performed on DBS samples in order to define the analytical performance of the TMA method starting from a minimum sample volume.

## Materials and methods

### Setting and study design

The University Hospital “Città della Salute e della Scienza di Torino” (Turin, Italy) is a regional tertiary hospital with 1900 beds spread over three structures: a general hospital, a pediatric clinic and a rehabilitation centre. The main area of excellence is represented by the transplantation of solid organs (mainly kidney and liver) and tissues (corneas, skin, skeletal muscle) in adult and pediatric patients.

The study was performed on a total of 755 whole blood samples collected in the period May-November 2022 at Microbiology and Virology Laboratory of A.O.U. “Città della Salute e della Scienza di Torino” for the diagnosis of HCMV infection or the monitoring of viremia in immunocompromised/transplanted patients. All samples were collected in K2/EDTA Vacutainer^®^ (Becton, Dickinson and Company, Franklin Lakes, USA). Epidemiological details of patients and distribution of wards involved are resumed in Table 1.

**Table 1.** Details of patients’ epidemiology and wards involved.

|  |  | Patients | Age |  |
| --- | --- | --- | --- | --- |
|  |  |  | mean | median |
|  | Male | 183 | 52 | 59 |
|  | Female | 140 | 48 | 54 |
|  | Ward | Patients | Samples |  |
|  | Kidney transplantation | 65 | 106 |  |
|  | HSC transplantation | 27 | 107 |  |
|  | Liver transplantation | 13 | 37 |  |
|  | Heart transplantation | 10 | 20 |  |
|  | Lung transplantation | 4 | 22 |  |
|  | Surgery | 38 | 91 |  |
|  | External facilities | 38 | 63 |  |
|  | Haematology | 26 | 44 |  |
|  | Nephrology | 14 | 33 |  |
|  | ICU/ED | 11 | 32 |  |
|  | Cardiology | 9 | 18 |  |
|  | Pneumology | 7 | 17 |  |
|  | Oncology | 7 | 1 |  |
|  | phlebotomy service | 6 | 9 |  |
|  | General medicine | 3 | 6 |  |
|  | Gastroenterology | 3 | 3 |  |
|  | Paediatric oncology | 13 | 58 |  |
|  | Paediatric transplantation centre | 8 | 33 |  |
|  | Paediatric nephrology | 7 | 8 |  |
|  | Paediatric cardiology | 4 | 14 |  |
|  | General paediatrics | 3 | 8 |  |
|  | Paediatric gastroenterology | 2 | 11 |  |
|  | Paediatric pneumology | 1 | 1 |  |
|  | Paediatric haematology | 1 | 1 |  |
|  | Rehabilitation centre | 3 | 12 |  |
|  | TOTAL | 323 | 755 |  |

### qPCR chemistry

The laboratory routine method (Standard of Care), used as reference in the current study, was a dual-step process performed as follow: an initial extraction of HCMV DNA from whole blood samples with DSP Mini Kit on QIAsymphony SP/AS platform (Qiagen^®^, Hilden, Germany) and subsequential amplification with CMV ELITe MGB^®^ qPCR kit (ELITechGroup, Turin, Italy) on ABI 7500 Fast Dx instrument (Applied Biosystems, Waltham, USA). Finally, results were converted in International Unit on ml (IU/ml) with the software Real Time System Analysis Software V. 2.0 (release 1.9991) (ELITechGroup, Turin, Italy). The tested viral region was the exon 4 region of the CMV MIEA gene (major immediate early antigen, HCMVUL123) with an amplicon size of 79 bp.

### TMA chemistry

Whole blood samples were diluted in a prefilled secondary tube containing the Aptima^®^ Whole Blood Diluent (Hologic Inc., Marlborough, USA) to a volume of 500 μl and 1425 μl, respectively. Then, each sample was loaded onto Panther system automated platform (Hologic Inc., Marlborough, USA) and tested with the Aptima CMV Quant assay kit (Hologic Inc., Marlborough, USA). The assay has three main steps: target capture, target amplification using TMA and detection of amplification products. Finally, the results were converted from copies/mL to IU/mL using a conversion factor equation embedded in the Panther software. For this kit, the viral region tested was the UL56 gene in the unique long (UL) region of the CMV genome with an amplicon size of 108 bp.

Both chemistries (qPCR and TMA) have been standardized to the WHO 1^st^ International Standard for human cytomegalovirus (NIBSC code: 09/162, United Kingdom).

### DBS

In order to evaluate the clinical performance of TMA chemistry on Dried Blood Spot (DBS), 52 whole blood positive samples with quantifiable viremia were selected and processed according to the following protocol: 50 μl of blood sample (approximately the volume of a drop) were spotted on Whatman protein saver™ 903™ cards (GE HealthCare, Marlborough, USA), dried for at least 24 hours at room temperature and stored at the same temperature for one week. Subsequently, each sample was pretreated with Aptima™ DBS Extraction Buffer (Hologic Inc., Marlborough, USA) according to the manufacturer’s instructions: briefly, 12 mm^3^ of DBS specimen was added to 1 ml of DBS Extraction Buffer and gently stirred at room temperature for 30 minutes. Then, vials containing the extracted DBS were centrifuged for 2 minutes at 3,000 g and loaded onto the Panther System. Each sample was tested in duplicate with Aptima CMV Quant assay kit.

### Clinical agreement

Whole blood samples were tested simultaneously with qPCR and TMA chemistries in order to evaluate overall sensitivity, specificity and quantification agreement. Moreover, 12 selected patients were monitored in terms of HCMV viraemia for up to three months of hospitalization; the inclusion criteria for patients’ selection were: (1) solid organ transplantation, (2) no less than four quantifications for at least 14 days, (3) HCMV viraemia detectable in the first blood sampling with at least one of the two tested methods.

### Statistical analysis

Data were analyzed as Log_10_-transformed values. In the correlation analysis, only samples in which the assays had quantitative values were considered. The correlation between the quantitative results was evaluated with linear regression analysis and Bland-Altman plot. The correlation coefficient was calculated using Pearson’s correlation. All analysis was performed using MedCalc statistical software Program version 20.305 (MedCalc Software Ltd, Ostend, Belgium).

## Results

TMA and qPCR performance data showed 99.28% agreement for quantified detected samples and 89.39% agreement for those negative between the two tested methods (Table 2). Cohen’s kappa coefficient was excellent (K-value: 0.87).

**Table 2.** Comparison of clinical agreement between TMA and qPCR assays. Compared to conventional qPCR, the TMA assay resulted in a higher number of detected or quantified samples (10.61%): 5 samples quantified with TMA chemistry had a mean value between 2.27 and 2.84 Log_10_ IU/ml, while one samples had a higher value of 3.92 Log_10_ IU/ml. When repeated, the result was confirmed. Additionally, for samples quantified with TMA assay and detected by qPCR, values ranged between 2.0 and 4.0 Log_10_ IU/ml, with the highest distribution for those 2.4 - 2.8 Log_10_ IU/ml (36/71, 50.70%) (Figure 1).

| qPCR | TMA |  |  | Total | Agreement (%) |
| --- | --- | --- | --- | --- | --- |
|  | Not detected | Detected | Quantified |  |  |
| Not detected | 337 | 34 | 6 | 377 | 89.39% |
| Detected | 9 | 23 | 71 | 103 | 22.33% |
| Quantified | 0 | 2 | 273 | 275 | 99.27% |
| Total | 346 | 59 | 350 | 755 |  |

**Figure 1.**
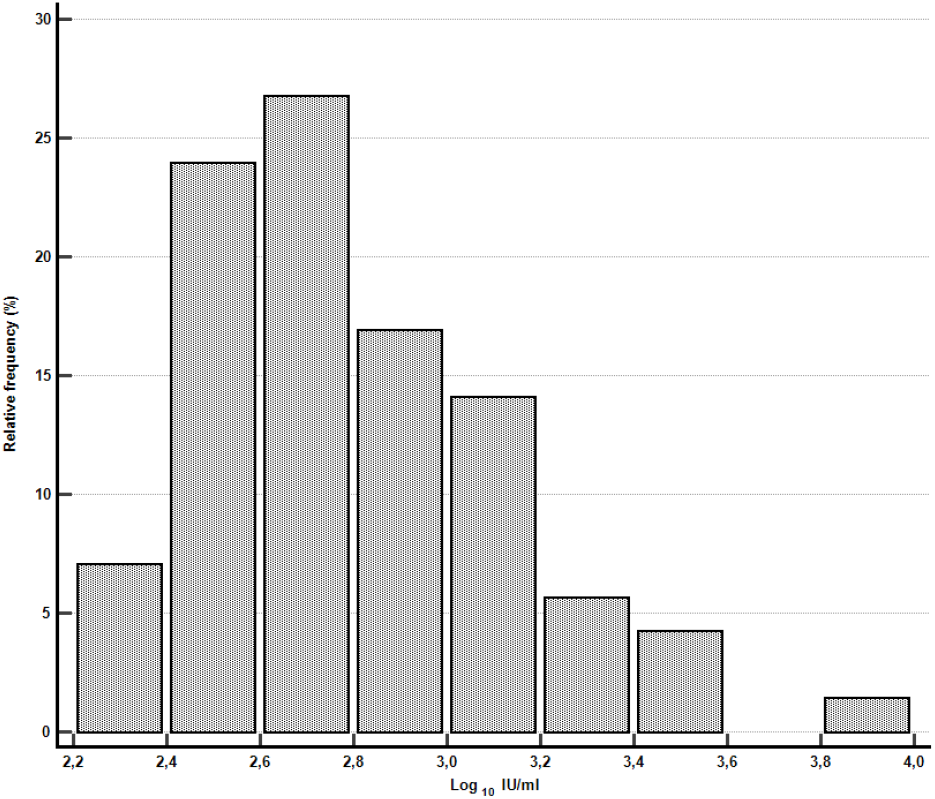
Distribution of Log_10_ IU/ml values with TMA assay in qPCR detected samples. Otherwise, 9 samples tested positive with qPCR and not detected with TMA, while two were quantified by qPCR assay only: the mean value was 2.65 ± 0.14 Log_10_ IU/ml (Table 1). A total of 273 samples had quantifiable results on both assays and were used for the correlation analysis. Deming’s regression analysis showed a significant correlation between the two chemistries: the linear regression line was calculated as follows, Y = 0.4581 + 0.8024 x with an R^2^ = 0.7142 (Figure 2). Pearson’s correlation coefficient value was 0.8796 (95% CI: 0.8496 to 0.9039) (P-value < 0.0001).

**Figure 2.**
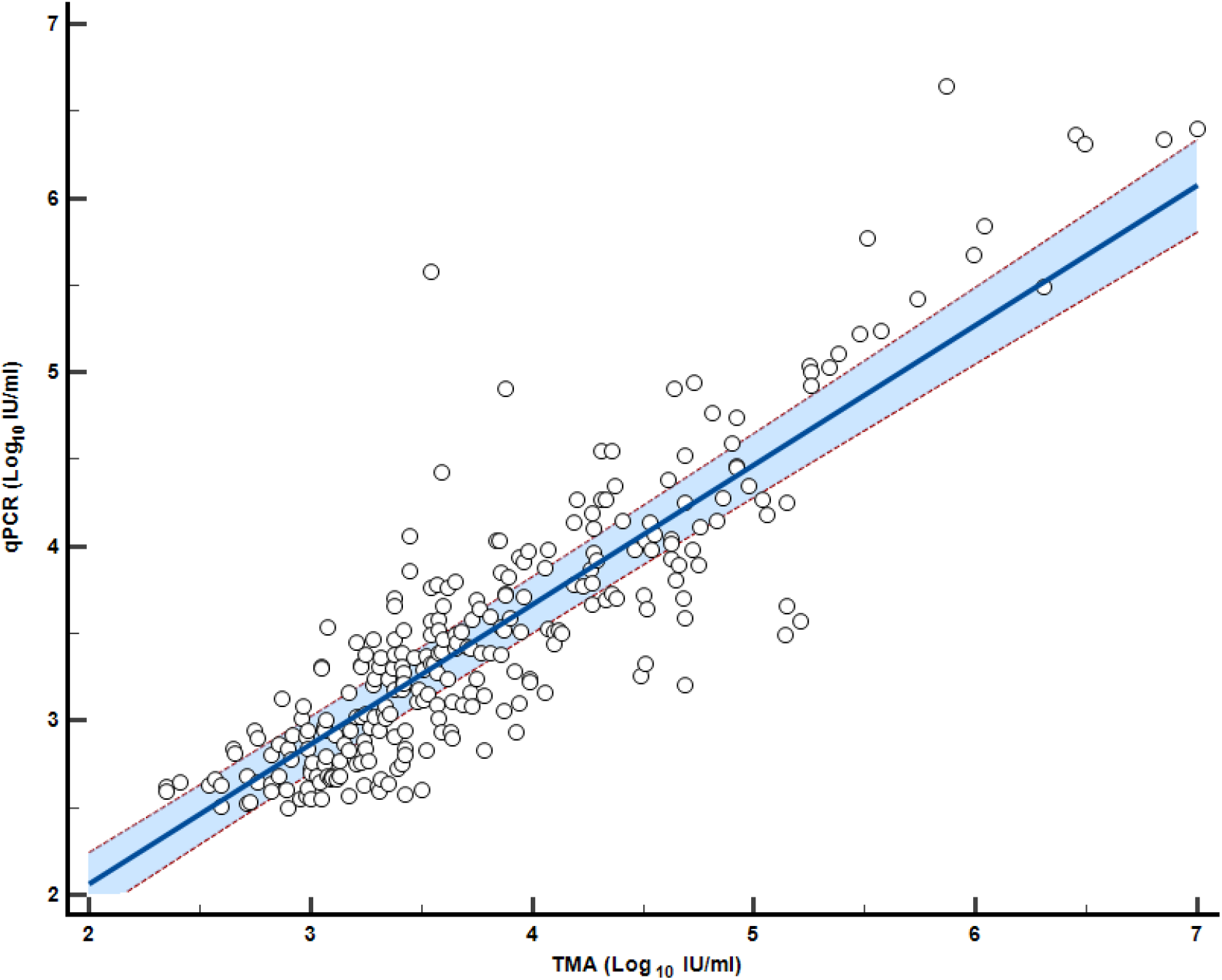
Log_10_-transformed quantitative viral load agreement for TMA and qPCR chemistry: linear regression. The mean viral load (±SD) of the TMA assay was 3.79 ± 0.83 Log_10_ IU/ml, while that obtained by qPCR was 3.50 ± 0.80 Log_10_ IU/ml. The Bland-Altman analysis (Figure 3) yielded a mean bias of −0.29 Log_10_ IU/ml (SD ± 0.40 Log_10_ IU/ml). The lower and upper limits of agreement were −1.07 (95% CI: −1.15 to −0.99) and 0.50 (95% CI: 0.42 to 0.58), respectively.

**Figure 3.**
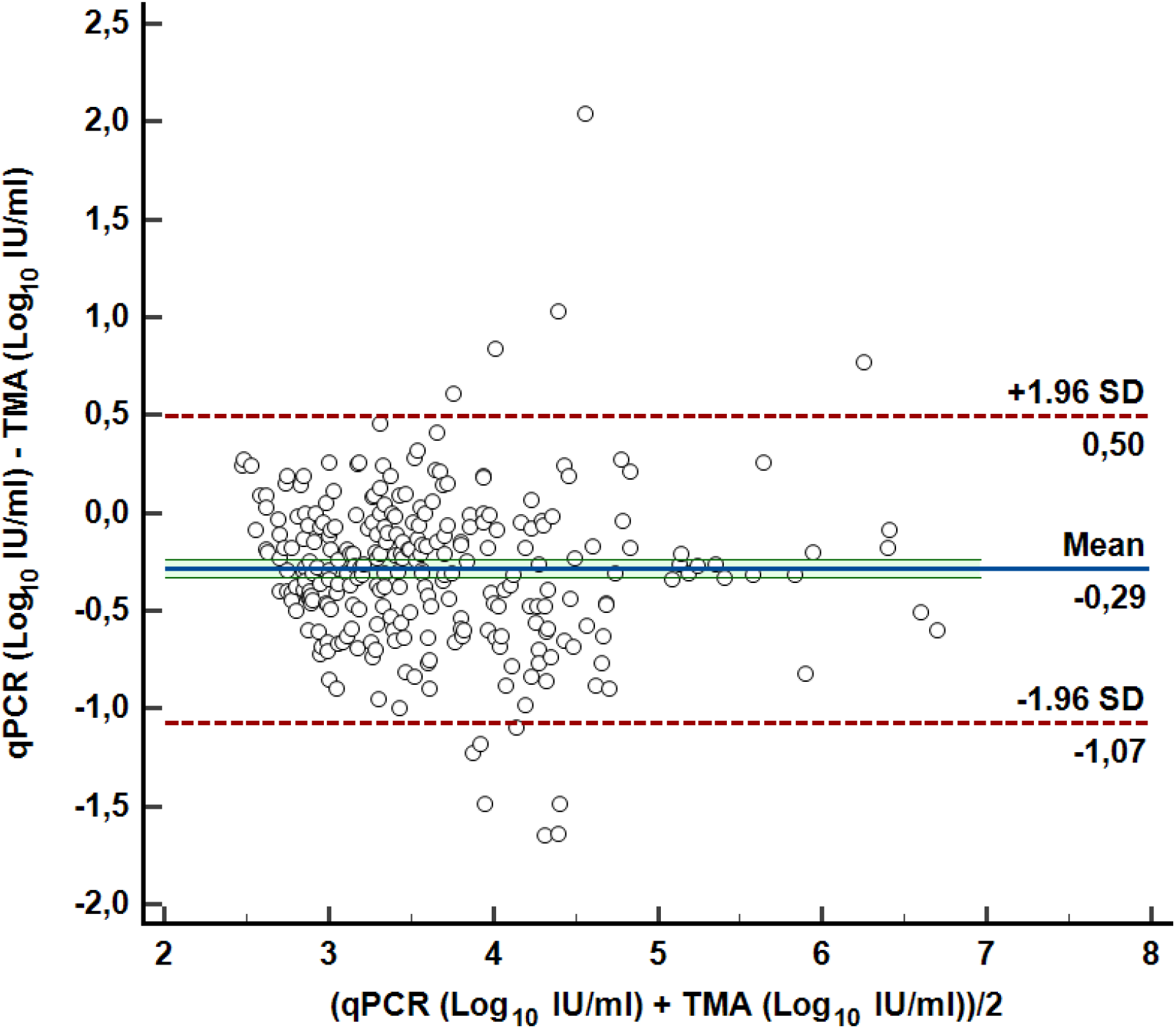
Log_10_-transformed quantitative viral load agreement for TMA and qPCR chemistry: Bland-Altman plot. According to the abovementioned criteria, 12 transplanted patients were monitored for a period ranging from 17 days to 100 days, depending on the hospital stay (Figure 4). All patients tested positive for both methods at time 0, except one (Figure 4, Patient #7) in which initial viremia was only detectable by TMA assay. Overall, in the samples tested from these patients, the mean difference between qPCR and TMA chemistries was − 0.59 ± 0.63 Log_10_ IU/ml. The strictest agreement resulted for patient number 5, 9, 10, 12 (mean difference − 0.23 ± 0.06 Log_10_ IU/ml). Interestingly, the patient #1 resulted steadily positive for up to three months of monitoring with TMA assay, while qPCR tested negative at day 17. However, the previously day, both assays resulted positive (2.48 or less and 3.45 Log_10_ IU/ml respectively for qPCR and TMA chemistry). When retested, the trend of HCMV viremia was confirmed in the samples from both days.

**Figure 4.**
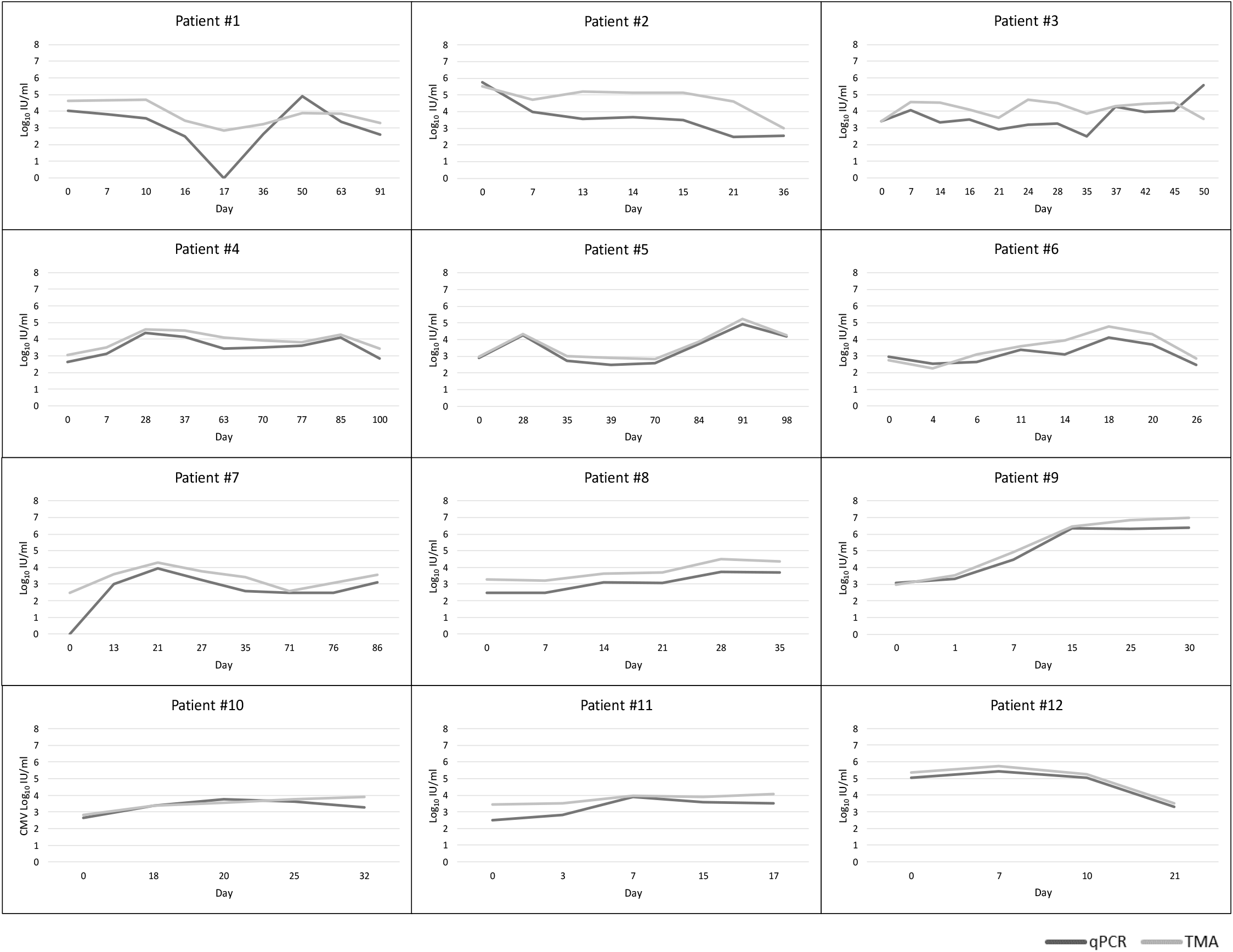
Trend of HCMV viremia with qPCR and TMA assays in transplanted patients. 52 quantified samples tested simultaneously as whole blood and spotted on DBS. Overall agreement was 86.54% (Table 3). 7 specimens (13.46%) resulted negative and, using a conversion factor of 36.4 (37), the mean value was 1.72 ± 0.54 Log_10_ IU/ml. Among positive results, 8 samples (15.38%) tested detected but not quantified on DBS: tough the conversion factor abovementioned, mean value for those specimens was 1.43 ± 0.42 Log_10_ IU/ml.

The most evident difference between tested assay resulted in patient #2 with a mean value of − 1.12 ± 0.84 Log_10_ IU/ml.

**Table 3.**
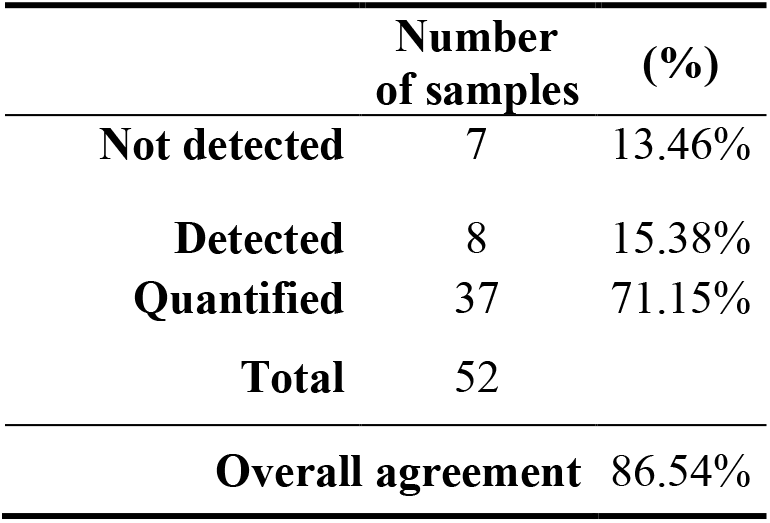
TMA assay: overall agreement between whole blood samples and spotted on DBS. Focusing on 37 quantified specimens both on whole blood and DBS, regression analysis using the aforesaid conversion factor showed a significant correlation between the two chemistries: the linear regression line was calculated as follows, Y = 1.4257 + 0.5936 x with an R^2^ = 0.7232 (Figure 5). Pearson’s correlation coefficient value was 0.8977 (95% CI: 0.8090 to 0.9464) (P-value < 0.0001).

**Figure 5.**
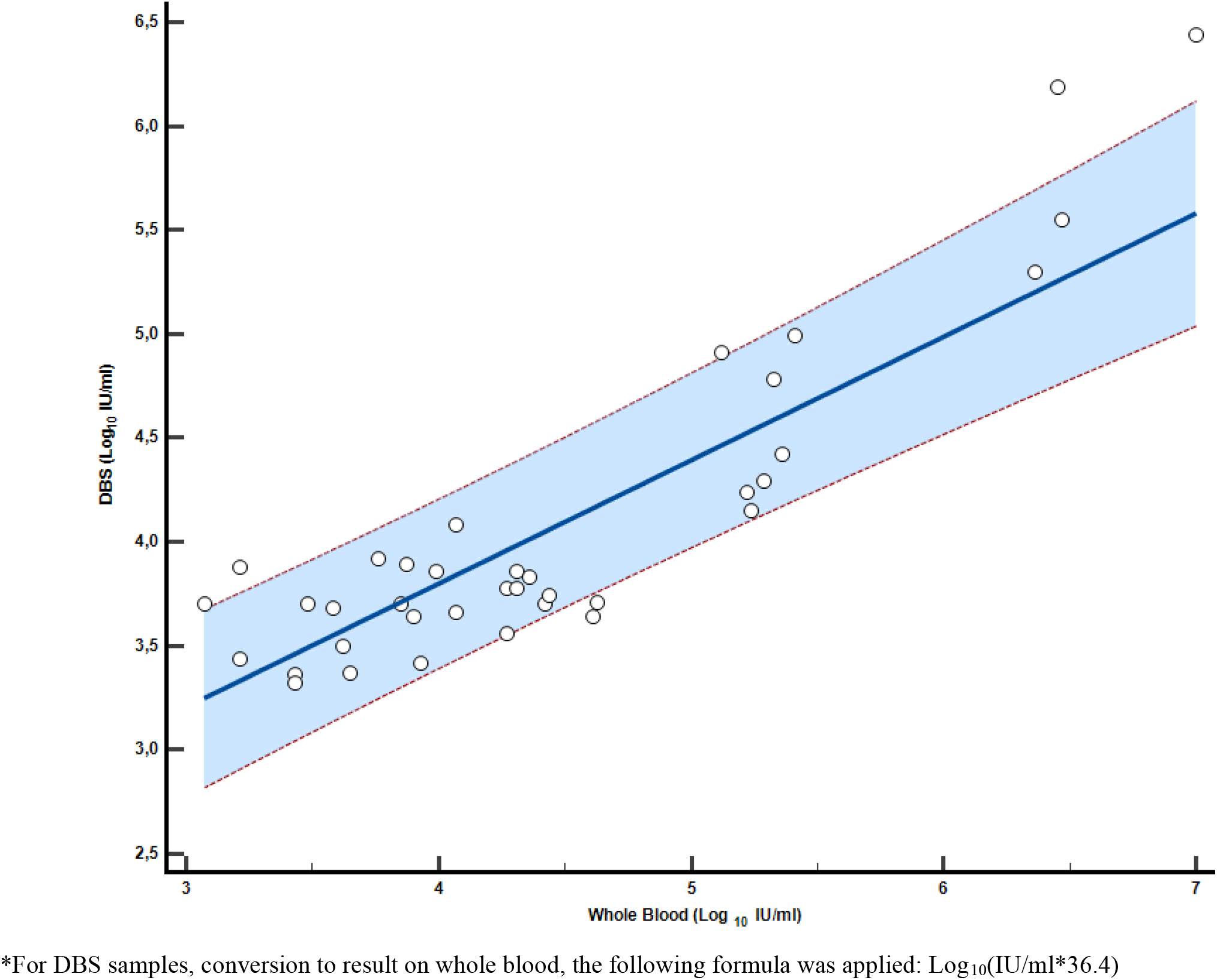
Log_10_-transformed quantitative viral load agreement for whole blood samples and the same spotted as DBS* with TMA assay: linear regression. *For DBS samples, conversion to result on whole blood, the following formula was applied: Log_10_(IU/ml*36.4)

## Discussion

Early detection of HCMV viraemia is pivotal to identify progressing infection and patients at high risk, which required antiviral therapy. Especially, the effectiveness of a pre-emptive therapy relies on accurate laboratory tests to monitor HCMV infection (38). Specimens currently recommended as the gold standard for monitoring the presence of HCMV DNA are whole blood and plasma that contain it respectively in monocytes and as free fragmented HCMV genome, which is released from infected cells (34). Although there is no consensus on the optimal blood compartment for routine molecular testing of HCMV DNA and the stability of the viral genome in both whole blood and plasma up to 14 at 4°C (39, 40), the higher sensitivity of whole blood and its higher HCMV DNA yield make it an optimal sample for monitoring viral load in immunocompromised patients (41). Moreover, in order to enhance interlaboratory agreement of HCMV DNA quantification, also in relation to the first WHO International Standard established in October 2010, different diagnostic workflows (extraction and amplification) were evaluated to define an appropriate conversion factor (CF) from genome/ml to IU/ml both on whole blood and plasma in a multicentric study (42, 43). Also due to this standardization, it has been possible to better understand viral kinetics and threshold values for predicting HCMV disease: although the viral load peak was reached simultaneously in the two blood compartments, HCMV viraemia in whole blood was constantly higher (about 1 log_10_ IU/ml) than in plasma since the onset of infection until the peak of viral load and it decreased more rapidly than in plasma, thus avoiding unnecessary prolonged treatment and saving in terms of drug toxicity and cost (44).

At the time of writing, several qPCR diagnostics assays have been developed and are currently available for identification and quantification of HCMV in plasma and whole blood; besides these, also a recently new TMA assay was introduced.

Differently from qPCR, target amplification via TMA is a transcription-mediated nucleic acid amplification method that uses two enzymes, Moloney murine leukemia virus (MMLV) reverse transcriptase and T7 RNA polymerase. The first enzyme generates a DNA copy of UL56 HCMV gene containing a promoter sequence for T7 RNA polymerase, which subsequently produces multiple copies of RNA amplicon from the viral DNA copy template. Detection is achieved using single-stranded nucleic acid torches, provide of a fluorophore and a quencher, that hybridize specifically to the amplicon of HCMV DNA.

Although correlation between qPCR and TMA chemistries for HCMV detection was assessed on plasma and other types of samples with excellent concordance (45, 46), EDTA/whole blood as clinical samples were not yet investigated.

In this study, we evaluated the performance of the TMA assay (Aptima^®^ CMV Quant Assay) and compared the data with our qPCR SoC (CMV ELITe MGB^®^ Kit). Clinical agreement between two chemistries was excellent (99.28% and 89.39% for quantified and negative samples, respectively). Moreover, TMA assay quantify at an average of 0.29 Log_10_ IU/mL higher compared to the qPCR assay, as showed in the Bland-Altman plot (Figure 3). Since mean difference was less than 0.5 Log, the cut-off to consider load changes biologically important (47), our results could suggest no clinical implication if diagnostic platform will be changed. However, this could be applied for our clinical setting where a side-to-side evaluation has been performed. Indeed, data about plasma samples showed a mean difference variable between – 0.20 and 0.58 Log_10_ IU/mL, according to TMA compared with different qPCR platform (45, 46).

Herein, TMA assay resulted more sensitive, resulting in a higher number of samples with a defined viral load (Table 2). This is probably due to the differences between the two methods in terms of Limit of Detection (LoD) and Limit of Quantification (LoQ). This latter amounted on 2.18 Log_10_ IU/ml for TMA assay and 2.48 Log_10_ IU/ml for qPCR; several discrepant samples stood between the range 2.2 - 2.8 Log_10_ IU/ml, thus near the lower LoQ of the qPCR assay (Figure 1). It is noteworthy that TMA assay needed a higher extraction volume (700 μl) but it must be diluted in ratio 1:2.85 with the appropriate preparation buffer, while for qPCR Starting volume is 200 μl of whole blood directly from vacutainer eluted in a final volume of 110 μl. Moreover, TMA assay was evaluated with a fully automated sample-to-result platform, while two different platforms, one for HCMV DNA extraction and the other for amplification, were used in combination for qPCR SoC. All abovementioned aspects, together with gene target and amplicon size (48), have to be taken into account to explain the differences between the two tested methods.

TMA chemistry performances were assessed to monitor viremia in transplanted patients; twelve individuals were selected and tested for a period of time with both assays. Our data showed a good overlay until more than three months, with TMA values on average higher than qPCR (Figure 4), in agreement with overall results. The major discrepancy was observed in patient 1, in which qPCR resulted suddenly not detected while TMA assay showed a linear trend in HCMV viraemia. Nevertheless, due to the reduced number of patients enrolled, no correlation with antiviral therapy and clinical outcome were evaluated. Another limitation of our study in this patients’ cohorts is the lackness of data about antiviral-resistant HCMV strains that could affect the compared methods.

For an in-depth analysis of whole blood as clinical specimen, TMA clinical performances were tested also with low volumes such as those obtained onto DBS. In this preliminary study, 52 positive whole blood samples were spotted onto DBS and results showed 86.54% of agreement (Table 3). For quantified specimens, applying the abovementioned conversion factor of 36.4, linear regression was excellent (Figure 5). Focusing on negative and detected DBS samples, mean viral load expected to be on whole blood was respectively 1.72 ± 0.54 Log_10_ IU/ml and 1.43 ± 0.42 Log_10_ IU/ml, thus under the LoD of TMA assay (2.18 Log_10_ IU/ml). Since low viral loads are unlikely to be associated to clinical consequences in cCMV infection (49), results herein reported can be considered acceptable for clinical use. To our knowledge, this is the first evaluation of TMA on low volumes resulted on Dried Blood Spot. However, the most important limit of this preliminary analysis is the need for a higher number of tested specimens obtained directly from patients, even for calculate a most suitable conversion factor between whole blood and DBS.

The excellent TMA testing results obtained in the present study, together with those of other studies, allow the opportunity of further studies to better understand the impact of TMA assay, especially in transplanted patients, on clinical management and decision to begin preemptive therapy or antiviral prophylaxis. However, due to its recent introduction for HCMV diagnosis, no data of comparison between plasma and whole blood are available to assess the most suitable matrix with TMA chemistry.

Alongside clinical and analytical performances, TMA assay showed the advantage to be carried out onto a fully automated system with random and continuous access, thus eliminating the need to batch samples and leading to faster turnaround time.

Laboratories evaluating a change to a different diagnostic system must considered all previously mentioned aspects performing a side-to-side testing using both current and new assays to understand the most suitable diagnostic workflow and clinical matrix.

TMA chemistry is a reliable diagnostic assay that allows accurate assessment of HCMV viremia on whole blood specimens, even with small volumes such as those of Guthrie card collected for diagnosis of cCMV infection. Moreover, this is the first TMA kit with FDA and CE-IVD approval on whole blood samples, thus clinical laboratories can easily incorporate this test in their routine molecular workflows. The availability of an automated platform with minimal hand-on time also enables timely diagnosis, prognosis and monitoring of HCMV infected patients.

## Data Availability

All data produced in the present study are available upon reasonable request to the authors

## Acknowledgment

This research was supported by EU funding within the MUR PNRR Extended Partnership initiative on Emerging Infectious Diseases (Project no. PE00000007, INF-ACT).

## Notes

### Competing Interest Statement

The authors have declared no competing interest.

### Funding Statement

This study did not receive any funding

### Author Declarations

The study was conducted in accordance with the Declaration of Helsinki, and approved by the Intercompany ethics committee of University Hospital Citta' della Salute e della Scienza di Torino, A.O. Ordine Mauriziano di Torino, A.S.L. Citta' di Torino (protocol code 0041695 of 15/04/2021).

